# The Causal Nexus of Immune Cells and Vitiligo: A Genetic Perspective

**DOI:** 10.1101/2024.11.03.24316683

**Authors:** Ziyi Lin, Zhimin Wu, Xingwu Duan

## Abstract

**Objective:** This study aims to explore the causal relationship between 731 immune cell traits and vitiligo using Mendelian randomization.

**Methods:** We used a two-sample Mendelian randomization (MR) analysis, employing genetic variations extracted from genome-wide association studies (GWAS) as instrumental variables (IVs). Data sources included immune phenotype data from 3,757 European individuals and data from FinnGen, comprising a total of 385,538 samples (292 cases and 385,509 controls). The study used the inverse-variance weighted (IVW) method as the primary analysis method and conducted various robustness tests through median-based weighted analysis, mode-based weighted analysis, and the MR-Egger method to control for false positive results in multiple hypothesis testing.

**Results:** Our study found that the pathogenesis of vitiligo may significantly reduce the levels of the following immune cells: TD CD4+ %T cells (b=-0.458, 95% CI=0.17-0.76, PFDR=0.015, P=0.000), CD25 on CD39+ CD4+ cells (b=-0.155, 95% CI=0.78-0.94, PFDR=0.166, P=0.769), and CD4 on HLA DR+ CD4+ cells (b=-0.431, 95% CI=0.50-0.85, PFDR=0.166, P=0.001). Additionally, the causal effect estimate of vitiligo on CCR2 on monocyte is 0.75 (b=-0.290, 95% CI=0.64-0.88, PFDR=0.114, P=0.828), and a negative association was also found on FSC-A on NK cells (b=-0.481, 95% CI=0.47-0.82, PFDR=0.142, P=0.197). On the other hand, our study suggests that the occurrence of vitiligo may increase the levels of CD28 on CD28+ CD45RA+ CD8br cells (b=0.311, 95% CI=1.13-1.65, PFDR=0.166, P=0.393).

**Conclusion:** The results of this study reveal causal links between vitiligo and multiple immune cell traits, highlighting the important role of the immune system in the pathogenesis of vitiligo. These findings provide new research directions for controlling vitiligo through immune regulation and may pave the way for early intervention and treatment strategies.

## 1 Introduction

Vitiligo is a chronic skin disease characterized by the loss of function of skin pigment cells, leading to the appearance of white patches on the skin (**Error! Reference source not found**.). The incidence of vitiligo is approximately 0.5% to 2%, affecting millions of people worldwide (**Error! Reference source not found**.). This disease can manifest at any age but usually appears before the age of 20. Vitiligo not only impacts the appearance of patients but can also have profound effects on their mental health, leading to decreased self-esteem, social barriers, and reduced quality of life (**Error! Reference source not found**.). Therefore, vitiligo is not only a medical issue but also a psychosocial problem, requiring comprehensive treatment and care to improve the patients’ quality of life.

While the exact pathogenesis of vitiligo has not been fully elucidated, increasing evidence suggests that the immune system plays a crucial role in its development. Laboratory findings indicate that the abnormal secretion of inflammatory markers such as TNF-α and interleukin-6 (IL-6) may significantly contribute to the pathogenesis of vitiligo. Additionally, the activation and functional changes of immune cells like monocytes and T cells can affect the survival and function of melanocytes, leading to the loss of skin pigmentation (**Error! Reference source not found**.). These studies suggest that regulating the immune system holds great potential in controlling vitiligo. However, it remains an area of exploration whether the persistent loss of skin pigmentation and immune system abnormalities in vitiligo patients are influenced by deeper genetic variations. Some scholars propose that acquired genetic and epigenetic risks play a significant role in explaining the different susceptibilities to vitiligo, considering its highly complex and polygenic nature (**Error! Reference source not found**.). To date, only a few risk loci for vitiligo have been identified. Furthermore, studies have found that vitiligo has familial heredity, focusing on candidate genes such as NLRP1, PTPN22, and TYR to explore the clinical genetics of vitiligo (**Error! Reference source not found**.). Research also aims to seek safer and more effective approaches through pharmacogenomics (PGx) to address the high non-responsiveness or partial responsiveness of vitiligo patients to existing treatments (**Error! Reference source not found**.). Genome-wide association studies (GWAS) play a key role in analyzing genetic variations across the entire genome in large populations, identifying expected genetic loci and pathways, and enhancing our understanding of the complex genetic factors of the disease (**Error! Reference source not found**.). This will also help to further explore the relationship between susceptibility, immune inflammation, and genetic background in vitiligo patients in our studies.

Mendelian randomization (MR) is a statistical method primarily based on the principles of Mendelian genetics to infer causal relationships in epidemiology (9). In the Mendelian randomization approach, ensuring the logical sequence of causal relationships is crucial (10). Previous observational studies (**Error! Reference source not found**.) have revealed certain associations between immune cell traits and vitiligo, supporting the hypothesis of their correlation. This study conducted a thorough two-sample MR analysis to explore the causal relationship between immune cell traits and vitiligo.

## 2 Materials and methods

### 2.1 Study design

We used a two-sample Mendelian randomization (MR) analysis method to study the causal association between 731 immune cell traits and generalized vitiligo. MR uses genetic variation as a substitute for risk variables and requires instrumental variables (IVs) to satisfy three key assumptions for causal inference: (1) the exposure is directly associated with the genetic variation; (2) there is no genetic association between exposure and outcome due to potential confounding factors; (3) there is no genetic effect on the outcome through pathways unrelated to the exposure (see Figure 1). Our study was authorized by the appropriate institutional review board, and informed consent was obtained from the participants.

**Fig. 1.**
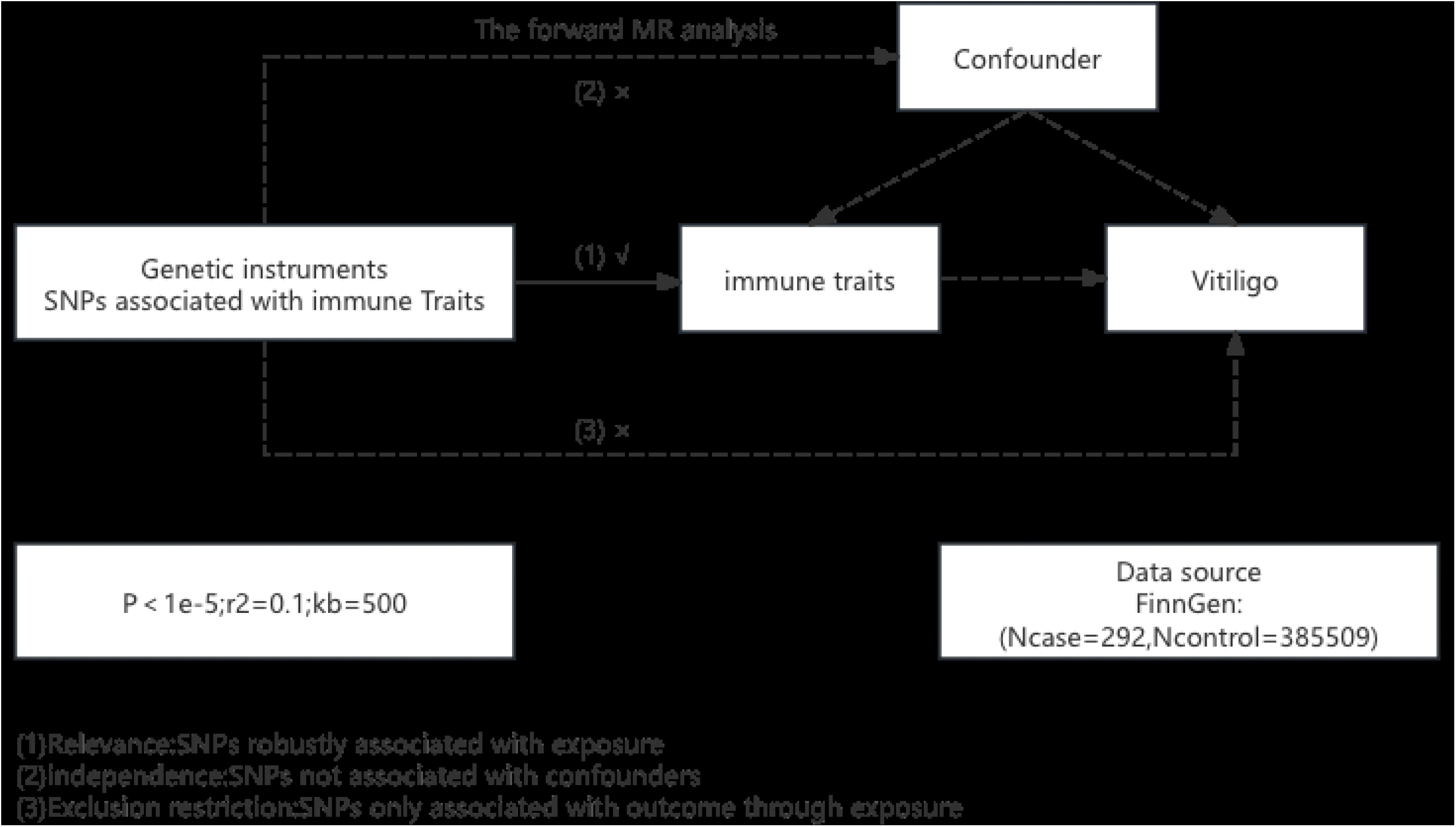

### 2.2 Sources of immunity-spanning GWAS data

The summary statistics for all immunological traits in the GWAS catalog (from accession number GCST0001391 to GCST0002121) are readily available (**Error! Reference source not found**.). The GWAS involved 3,757 non-overlapping European individuals. Based on a Sardinian sequence reference panel, a high-density array of approximately 22 million single nucleotide polymorphisms (SNPs) was estimated, and correlation tests were conducted after controlling for covariates such as age, age squared, and sex (13). A total of 731 immune phenotypes were examined, including relative cell counts (RC) (192), morphological parameters (MP) (32), absolute cell counts (AC) (118), and median fluorescence intensity (MFI) (389) representing surface antigen levels. Among them, MP traits included CDC and TBNK panels, while MFI, RC, and AC traits included B cells, CDC, T cell maturation stages, myeloid cells, monocytes, and TBNK (T cells, B cells, natural killer proteins).

### 2.3 Data sources from the genome-wide association study for GAD

The summary statistics for the genome-wide association study (GWAS) of vitiligo were obtained from FinnGen (https://www.finngen.fi/en). The study covered a total of 385,538 samples (cases = 292, controls = 385,509), with a median age of 45.91 years at the first occurrence of vitiligo (42.35 years for females, 48.21 years for males). The GWAS analysis included 470,000 phenotype data points related to vitiligo, identifying over 21.31 million independent single nucleotide polymorphisms (SNPs).

In all studies, R software version 4.4.1 (http://www.Rproject.org) was used. To specifically assess the causal relationship between 731 immune phenotypes and vitiligo, we used the “MendelianRandomization” software (version 0.6.6) and conducted median-based weighted analysis, mode-based weighted analysis, and inverse-variance weighted (IVW) analysis. Cochran’s Q statistic and its p-value (IV) were used to evaluate instrument heterogeneity among variables, and the MR-Egger method was employed to identify horizontal pleiotropy; if the intercept term was significant, horizontal pleiotropy was considered present. Finally, we used funnel plots, scatter plots and leave-one-out analysis. The scatter plots showed minimal impact of outliers on the data, while the funnel plots demonstrated high correlation and a lack of heterogeneity.

### 2.4 Nstrumental variables

A set of instrumental variables (IVs) (version v1.90) was selected (linkage disequilibrium [LD] r^2^ threshold less than 0.1 within a 500 kb distance) to modify these single nucleotide polymorphisms (SNPs) (14). The 1000 Genomes Project was used as the reference panel for calculating LD r^2^. A new significance threshold for vitiligo was set at 5 × 10^−8^. To evaluate the strength of the IVs and reduce the bias from weak instruments, the F-statistic was calculated.Statistical analysis.

## 3 Results

### 3.1 Examination of the causal relation of vitiligo onset on immunophenotypes

In exploring the causal effects of vitiligo on immune phenotypes, we used the inverse-variance weighted (IVW) method in two-sample Mendelian randomization (MR) analysis as the primary analysis method. After multiple testing adjustments based on the false discovery rate (FDR) method, seven suggestive immune phenotypes were identified at a significance level of 0.20, with four in the T cell group, one in the NK cell group, and one in the monocyte group.

Our study results suggest that the pathogenesis of vitiligo may significantly reduce the levels of the following immune cells: TD CD4+ %T cells (b=-0.458, 95% CI=0.17-0.76, PFDR=0.015, P=0.000), CD25 on CD39+ CD4+ cells (b=-0.155, 95% CI=0.78-0.94, PFDR=0.166, P=0.769), and CD4 on HLA DR+ CD4+ cells (b=-0.431, 95% CI=0.50-0.85, PFDR=0.166, P=0.001). Additionally, the causal effect estimate of vitiligo on CCR2 on monocyte is 0.75 (b=-0.290, 95% CI=0.64-0.88, PFDR=0.114, P=0.828), and a negative association was also found on FSC-A on NK cells (b=-0.481, 95% CI=0.47-0.82, PFDR=0.142, P=0.197).

On the other hand, our study suggests that the occurrence of vitiligo may increase the levels of CD28 on CD28+ CD45RA+ CD8br cells (b=0.311, 95% CI=1.13-1.65, PFDR=0.166, P=0.393) and the levels of CD8 on EM CD8br (b=0.378, 95% CI=1.19-1.79, PFDR=0.091, P=0.021).

The results of the other three methods and sensitivity analyses confirmed the robustness of the observed causal associations (Supplementary Table 1). Specifically, the global test of the MR-Egger intercept ruled out the possibility of horizontal pleiotropy (Supplementary Table 1). Scatter plots, funnel plots and leave-one-out analysis are contented in Supplementary Materials.

### 3.2 Sensitivity analysis

The results of the Cochran Q test indicated no heterogeneity among the SNPs (Supplementary Table 1). The MR-Egger intercept test results showed that horizontal pleiotropy did not affect the MR analysis outcomes (Supplementary Table 1). The funnel plot displayed a symmetrical distribution on both sides, suggesting no bias in the MR analysis results (Supplementary Figure 2). Sensitivity analysis using the leave-one-out method demonstrated that the MR analysis results were not influenced by individual SNPs (Supplementary Figure 3).

## 4 Discussion

We utilized a large amount of publicly available genetic data to investigate the causal relationship between 731 immune cell traits and vitiligo. To date, this is the only MR (Mendelian Randomization) study that has examined the causal link between 731 immune phenotypes and vitiligo. The study included four categories of immune traits (MFI, RC, AC, and MP). Among them, vitiligo showed causal effects on seven immune phenotypes (PFDR<0.20), and one immune phenotype had a strong causal effect on vitiligo (PFDR<0.05).

Our study suggests that the pathogenesis of vitiligo may reduce the levels of TD CD4+ %T cells, CD25 on CD39+ CD4+ cells, and CD4 on HLA DR+ CD4+ cells. CD4+ T cells primarily maintain normal immune function in the body. When the quantity and function of T lymphocyte subpopulations become abnormal, it can lead to immune dysfunction and a series of pathological changes (**Error! Reference source not found**.). In the development of vitiligo, CD4+ T cells can recognize exogenous antigen peptides and differentiate into effector cells, mainly Th cells. Upon activation, Th cells secrete large amounts of cytokines such as interleukin-2, interleukin-6, granulocyte-macrophage colony-stimulating factor, and tumor necrosis factor-α. These cytokines influence melanocytes by regulating innate and adaptive immunity or by directly inducing apoptosis, leading to the destruction of melanocytes and the onset of vitiligo. Research by Czarnowicki (**Error! Reference source not found**.) found that the expression of CD4+ T lymphocyte subpopulations in the peripheral blood of vitiligo patients is significantly lower than that of healthy individuals. Additionally, research by Dwivedi (**Error! Reference source not found**.) and Nigam (**Error! Reference source not found**.) also found that the proportion of CD3+ T cells and CD8+ T cells in the peripheral blood of vitiligo patients is increased, while the proportion of CD4+ T cells and the CD4+/CD8+ T cell ratio is decreased.

The expression of CCR2 on monocytes is closely associated with the pathogenesis of chronic skin diseases. To date, research on the role of CCR2 in skin diseases is limited. A clinical study found an abnormal increase of CCR2-positive monocytes in the peripheral blood of patients with atopic dermatitis and psoriasis, which is closely related to the immune dysregulation in these patients (**Error! Reference source not found**.). Mechanistically, CCR2-positive monocytes may promote the onset and progression of vitiligo by participating in the regulation of immune responses and influencing the survival and function of melanocytes (**Error! Reference source not found**.).

Our research found that the occurrence of vitiligo is associated with an increase in CD28+ CD45RA+ CD8br T cells and CD8 on EM CD8br cells. Van den Boorn and colleagues confirmed that, following immune system abnormalities, cytotoxic CD8+ T cells, aided by signaling factors, target and damage melanocytes, leading to the loss of their ability to normally produce melanin. This results in the depigmented lesions observed in the epidermis and hair of vitiligo patients, which appear as milky white patches (**Error! Reference source not found**.). Numerous studies (**Error! Reference source not found**.) have shown that a large number of active cytotoxic CD8+ T cells infiltrate the lesion sites in most vitiligo patients.

It is noteworthy that in this study, a negative correlation was also found between the expression of FSC-A on natural killer (NK) cells and vitiligo. NK cells, as key initiators of adaptive immune responses, are involved in the early development of vitiligo. NK cells, along with type 1 innate lymphoid cells (ILC-1), are part of the innate lymphoid cell family, forming the first line of defense against infection triggers and tumor cells (**Error! Reference source not found**.). Moreover, NK cells possess both cytotoxic and regulatory functions, capable of recognizing and destroying stressed cells through activating receptors, or limiting immune responses via inhibitory receptors (**Error! Reference source not found**.). Previous studies have reported elevated levels of circulating NK cells in vitiligo patients (**Error! Reference source not found**.), a finding that is not consistent with the results of this study. However, until recently, their role in the development of vitiligo remained unclear (**Error! Reference source not found**.). The association between NK cells and vitiligo still requires further investigation.

This study utilized a two-sample Mendelian randomization analysis, using data from a large genomic study cohort of approximately 385,538 individuals, ensuring high statistical efficiency. The results of the study were based on genetic instrumental variables and causal inference was conducted through various robust Mendelian randomization analysis techniques, which are not affected by horizontal pleiotropy and other variables. Additionally, to control for false positive results in multiple hypothesis testing, we employed the false discovery rate (FDR) to control for statistical bias due to multiple comparisons.

However, this study does have several limitations. First, this study did not employ bidirectional MR to analyze reverse causality. Second, due to the lack of individual-level data, stratified population analysis is not feasible. Third, the study relies on European databases, which limits the generalizability of the results to other racial groups. Finally, the flexible criteria for evaluating study outcomes may lead to more false positives, but they also facilitate a comprehensive assessment of the strong relationships between immune traits and vitiligo. Overall, the next step in the research on vitiligo mechanisms will be to reduce the impact of confounding factors to obtain higher levels of causal evidence.

## 5. Conclusion

In conclusion, our two-sample Mendelian randomization (MR) analysis revealed causal links between various immune phenotypes and vitiligo, elucidating the intricate relationship between vitiligo and the immune system. Additionally, the influence of other variables and unavoidable confounding factors in our study was successfully minimized, providing researchers with new perspectives to explore the biological basis of vitiligo and potentially paving the way for early intervention and treatment strategies. These findings extend the field of dermatology and offer valuable insights for the prevention of vitiligo.

## Data availability statement

The original contributions presented in the study are included in the article/Supplementary Material. Further inquiries can be directed to the corresponding author.

## Ethics statement

The data for this study were obtained from publicly accessible databases, thereby eliminating the need for further ethical approval. The studies were conducted in accordance with the local legislation and institutional requirements. Written informed consent for participation was not required from the participants or the participants’ legal guardians/next of kin in accordance with the national legislation and institutional requirements.

## Author contributions

ZL: Data curation, Formal analysis, Methodology, Resources, Software, Writing - original draft, Writing - review & editing. ZW: Conceptualization, Project administration, Supervision, Validation, Writing - review & editing. XD: Conceptualization, Funding acquisition, Project administration, Supervision, Validation, Visualization, Writing - review & editing.

## Funding

The author(s) declare financial support was received for the research, authorship, and/or publication of this article. This study was financially supported by Beijing University of Chinese Medicine Dongzhimen Hospital Clinical Research and Achievement Transformation Capability Enhancement Pilot Project(DZMG-XZYY-23009).

## Conflict of interest

The authors declare that the research was conducted in the absence of any commercial or financial relationships that could be construed as a potential conflict of interest.

The handling editor declared a shared parent affiliation, though no other collaboration, with several of the authors ZL, ZW, XD,at the time of the review.

## Publisher’s note

All claims expressed in this article are solely those of the authors and do not necessarily represent those of their affiliated organizations, or those of the publisher, the editors and the reviewers. Any product that may be evaluated in this article, or claim that may be made by its manufacturer, is not guaranteed or endorsed by the publisher.

---

**Table S1.**

| Exposure | Outcome | Method | nSNP | P-value | PFDR |  | OR(95%CI) |
| --- | --- | --- | --- | --- | --- | --- | --- |
| TD CD4+ %T cell | VITILIGO | MR Egger | 47 | 0.010 | 0.015 |  | 0.350(0.170 to 0.760) |
|  | VITILIGO | Weighted median | 47 | 0.002 |  |  | 0.590(0.420 to 0.820) |
|  | VITILIGO | Inverse variance weighted | 47 | 0.000 |  |  | 0.630(0.510 to 0.780) |
|  | VITILIGO | Simple mode | 47 | 0.150 |  |  | 0.590(0.290 to 1.200) |
|  | VITILIGO | Weighted mode | 47 | 0.015 |  |  | 0.550(0.340 to 0.870) |
| CD28 on CD28+ CD45RA+ CD8br | VITILIGO | MR Egger | 29 | 0.288 | 0.166 |  | 1.320(0.800 to 2.180) |
|  | VITILIGO | Weighted median | 29 | 0.053 |  |  | 1.330(1.000 to 1.770) |
|  | VITILIGO | Inverse variance weighted | 29 | 0.002 |  |  | 1.370(1.130 to 1.650) |
|  | VITILIGO | Simple mode | 29 | 0.202 |  |  | 1.370(0.850 to 2.190) |
|  | VITILIGO | Weighted mode | 29 | 0.033 |  |  | 1.400(1.040 to 1.880) |
| CD4 on HLA DR+ CD4+ | VITILIGO | MR Egger | 36 | 0.836 | 0.166 |  | 0.930(0.460 to 1.860) |
|  | VITILIGO | Weighted median | 36 | 0.109 |  |  | 0.760(0.540 to 1.060) |
|  | VITILIGO | Inverse variance weighted | 36 | 0.002 |  |  | 0.650(0.500 to 0.850) |
|  | VITILIGO | Simple mode | 36 | 0.380 |  |  | 0.750(0.400 to 1.420) |
|  | VITILIGO | Weighted mode | 36 | 0.339 |  |  | 0.770(0.460 to 1.300) |
| CD25 on CD39+ CD4+ | VITILIGO | MR Egger | 98 | 0.204 | 0.166 |  | 0.890(0.740 to 1.070) |
|  | VITILIGO | Weighted median | 98 | 0.083 |  |  | 0.870(0.750 to 1.020) |
|  | VITILIGO | Inverse variance weighted | 98 | 0.001 |  |  | 0.860(0.780 to 0.940) |
|  | VITILIGO | Simple mode | 98 | 0.432 |  |  | 0.890(0.680 to 1.180) |
|  | VITILIGO | Weighted mode | 98 | 0.051 |  |  | 0.850(0.720 to 1.000) |
| FSC-A on NK | VITILIGO | MR Egger | 22 | 0.025 | 0.142 |  | 0.320(0.130 to 0.800) |
|  | VITILIGO | Weighted median | 22 | 0.003 |  |  | 0.530(0.350 to 0.810) |
|  | VITILIGO | Inverse variance weighted | 22 | 0.001 |  |  | 0.620(0.470 to 0.820) |
|  | VITILIGO | Simple mode | 22 | 0.655 |  |  | 0.840(0.390 to 1.810) |
|  | VITILIGO | Weighted mode | 22 | 0.034 |  |  | 0.470(0.250 to 0.900) |
| CCR2 on monocyte | VITILIGO | MR Egger | 40 | 0.240 | 0.114 |  | 0.780(0.520 to 1.170) |
|  | VITILIGO | Weighted median | 40 | 0.014 |  |  | 0.740(0.580 to 0.940) |
|  | VITILIGO | Inverse variance weighted | 40 | 0.000 |  |  | 0.750(0.640 to 0.880) |
|  | VITILIGO | Simple mode | 40 | 0.265 |  |  | 0.770(0.490 to 1.210) |
|  | VITILIGO | Weighted mode | 40 | 0.115 |  |  | 0.740(0.520 to 1.070) |
| CD8 on EM CD8br | VITILIGO | MR Egger | 40 | 0.041 | 0.091 |  | 1.680(1.040 to 2.710) |
|  | VITILIGO | Weighted median | 40 | 0.066 |  |  | 1.300(0.980 to 1.720) |
|  | VITILIGO | Inverse variance weighted | 40 | 0.000 |  |  | 1.460(1.190 to 1.700) |
|  | VITILIGO | Simple mode | 40 | 0.570 |  |  | 1.200(0.650 to 2.210) |
|  | VITILIGO | Weighted mode | 40 | 0.516 |  |  | 1.150(0.760 to 1.730) |

*P*<0.05 was considered statistically significant

## Notes

### Competing Interest Statement

The authors have declared no competing interest.

### Funding Statement

Yes

